# Grandparental Childcare During the Pandemic and Effects on Mental Health: Evidence From England

**DOI:** 10.1101/2022.02.09.22270740

**Authors:** Giorgio Di Gessa, Valeria Bordone, Bruno Arpino

**Author notes:** **Corresponding Author:** Dr. Giorgio DI GESSA, PhD.

## Abstract

**Objectives:** Policies aiming at reducing rates of hospitalisation and death from COVID-19 encouraged older people to reduce physical interactions. For grandparents in England, this meant that provision of care for grandchildren was allowed only under very limited circumstances. Evidence also suggests that reduced face-to-face interactions took a toll on mental health during the pandemic. This study aims to investigate whether changes in grandchild care provision during the pandemic impacted grandparents’ mental health.

**Methods:** Using pre-pandemic data from Wave 9 (2018/19) and the second Covid-19 sub-study (November/December 2020) of the English Longitudinal Study of Ageing, we first describe changes in grandparenting since the start of the pandemic to then investigate, using regression models, associations between changes in grandparenting and mental health (depression, quality of life, life satisfaction) during the pandemic, while controlling for pre-pandemic levels of the outcome variables. Results: About 10% of grandparents stopped altogether to look after grandchildren during the pandemic, with 22% reporting an overall decrease in the amount of grandchild care provided and 20% an increase or similar levels. Compared to grandparents who mostly maintained unchanged their grandchild care provision, those who stopped altogether and those who mostly reduced the amount of grandchild care provided were more likely to report poorer mental health, even accounting for pre-pandemic health.

**Discussion:** While measures to limit physical contact and shield older people were necessary to reduce the spread of COVID-19, policymakers should acknowledge potential adverse consequences for mental health among grandparents who experienced changes in their roles as grandchild caregivers.

## Introduction

Early in the COVID-19 pandemic, it was shown that risks of serious illness and death deriving from COVID-19 increased with age and that many pre-existing diseases, also strongly correlated with age, increased morbidity and mortality risks (Guan et al. 2020; Jordan et al. 2020). Therefore, when governments enacted drastic public health measures to slow the spread of the virus and help prevent health services from being overrun (including lockdowns, shielding, and stay-at-home orders), there were clear recommendations that particularly older people should stay indoors, limit their travels and movements, as well as limit physical interactions with others (Ayalon 2020; Perra 2021). The implementation of such policies meant also the temporary suspension of grandchild care provision for many grandparents (Cantillon et al. 2021). The pandemic and associated policies restricting physical contact have however also posed a risk to mental health, with ample evidence suggesting that globally the COVID-19 pandemic has resulted in poorer mental health (Santomauro et al. 2021).

Although around the world grandparents as providers of childcare are a common and significant form of intergenerational family support (Di Gessa et al. 2020; Hank and Buber 2009; Hank et al. 2018; Ko and Hank 2014; Zamberletti et al. 2018), and human relationships, intergenerational family contacts as well as the provision of grandchild care itself are important factors for mental health (Danielsbacka et al. 2022; Nyqvist et al. 2013; Umberson and Montez 2010), to date no studies have investigated the impact of (changes in) grandparenting on mental health during the time of the COVID-19 pandemic. The present study, therefore, aims to provide new and robust evidence on the impact that changes in the provision of grandparental childcare during the pandemic had on grandparents’ mental health. Using data from the nationally-representative English Longitudinal Study of Ageing (ELSA), we investigate the associations between changes in the amount of time grandparents looked after grandchildren in the first 8-9 months of the pandemic and three important indicators of mental health (depression, life satisfaction, and quality of life), while accounting for socio-economic and demographic characteristics as well as pre-pandemic measures of mental health.

### Background

Before the COVID-19 Pandemic, around the world, grandparents as providers of childcare were a common and significant form of intergenerational family support. Although the definition of grandchild care and therefore its prevalence and intensity vary across studies and countries, a considerable body of work showed that grandparents were significant providers of secondary grandchild care (Di Gessa et al. 2020; Hank and Buber 2009; Hank et al. 2018; Ko and Hank 2014; Zamberletti et al. 2018). For instance, figures pre-pandemic show that more than 50% of grandparents looked after grandchildren in England (Di Gessa et al. 2020) and 20% of Italian grandchildren aged 0–13 were looked after by grandparents almost daily when their parents are at work (Zamberletti et al. 2018).

Especially in the early phases of the pandemic, as age is the most important factor in diminishing one’s chances to survive the COVID-19, governments’ strategy around the world has notably focused on targeting older people who have been advised not only to stay indoors and limit their interactions with others but also to stay away from their grandchildren and from younger people (Ayalon 2020; Perra 2021). Just to cite a few examples, the Israeli Ministry of Defence, Mr Bennett, stated that “the single most important insight is to separate old people from young people. The single most lethal combination cocktail is when grandma meets her grandchild and hugs him (Ayalon 2020).” In the UK, Prime Minister Mr Johnson stressed that grandparents should not look after children amid the coronavirus outbreak, with young people urged by the health secretary Hancock not to “kill [their] gran” (Rosney 2020). Also, several paediatricians, epidemiologists, and virologists suggested that overall “to be safe, grandparents really shouldn’t be doing childcare” (Glazer 2020).

While it is important to recognise that the clear aim and main benefits of guidelines focussing on staying at home and physical distancing are to contain the spread of the disease and save lives, such measures also all have the potential to substantially affect the mental health of the population both directly and indirectly (e.g. (Brooks et al. 2020; Pfefferbaum and North 2020)). The restriction of physical interactions has clearly posed a risk to mental health, and research investigating mental health sequelae of the pandemic both among older people (disproportionately impacted by shielding policies and stay-at-home advice) and in the general population has found substantial deterioration in mental health especially among those with pre-existing mental or physical health conditions and low social support (Armitage and Nellums 2020; Arpino et al. 2021; Brooks et al. 2020; Daly et al. 2020; Di Gessa and Price 2021a; Di Gessa and Price 2021b; Holmes et al. 2020; Pierce et al. 2020; Steptoe and Di Gessa 2021). However, little is known so far on the impact of the pandemic on grandparents who looked after grandchildren.

Although grandchild care provision per se might not explain the health differential observed between grandparents who look after grandchildren and those who do not (Ates 2017; Danielsbacka et al. 2019), and factors such as intensity and health outcomes add to the complexity of the relationship, several studies have found that grandparents providing grandchild care tend to report better health (Chen and Liu 2012; Di Gessa et al. 2016b, 2016c; Hughes et al. 2007; Ku et al. 2013). Positive associations have often been discussed in light of the emotional rewards and gratification stemming from this activity; the closeness of the relationships with grandchildren; stronger family ties and greater family support; and what is, to some, the most valued and satisfying of family roles (Danielsbacka et al. 2022; Hendricks 1995; Kivnick 1982). Therefore, abrupt changes or disruptions in intergenerational relationships might have negative consequences for the mental health of grandparents, particularly among those who suddenly stop looking after grandchildren. Previous studies that have investigated the consequences to the emotional well-being of grandparents after the loss of contact with their grandchildren (e.g., because of divorce or migration) suggest that such loss of contacts adversely affect grandparents’ mental health (including mental health problems, lowered life satisfaction, and depression), particularly when the loss was unexpected as well as when grandparents were unable to exert control over the situation and were longing for a get-together with their grandchildren (Drew and Smith 1999; Drew and Silverstein 2007). In addition to suffering from the lack of contact with their grandchildren, these grandparents also suffered from the loss of their grandparental role, the successful enactment of which has been linked to higher life satisfaction and well-being (Kivnick 1982; Reitzes and Mutran 2004). Although previous studies have suggested that non-physical contacts during the lockdown and the pandemic played a key role in attenuating the negative effects of the stressful context of COVID-19 on mental health (Arpino et al. 2021), we still expect that grandparents who during the pandemic stopped altogether to look after grandchildren or experienced significant disruptions to their role as grandchild carers might have experienced negative mental health consequences. This is in line with the concept of ‘ambiguous loss’ (Boss 2009) that, adapted to disruptions in the grandparent-grandchild relationship, theorises that the unfulfilled expectations and continued hope of reunion with their grandchildren can be a source of distress and emotional pain for grandparents who are not able to see and spend time with their grandchildren for reasons beyond their control.

In light of this, the present study aims to understand whether and to what extent disruptions in grandparent-grandchild relationships in the form of grandchild care provision impacted grandparents’ mental health during the pandemic (namely, depression, life satisfaction, and quality of life). To our knowledge, our study is the first to take into account changes in grandparental childcare provision during the first 8/9 months of the pandemic in England –exploiting newly collected data in England (see below). Moreover, given that older people who stayed at home, shielded, and minimised face-to-face contacts are also disproportionately likely to have had poorer health prior to the pandemic, in this study we account for pre-pandemic characteristics which could potentially lead to a higher risk of social isolation and therefore to deteriorating mental health. Finally, given the detrimental independent role of lack of non-physical contacts, social isolation, and loneliness on mental health and well-being, we also accounted for these important factors to better understand the effect of changes in grandparental childcare provision on grandparents’ mental health.

## Methods

### Study Population

We used data from the English Longitudinal Study of Ageing (ELSA) (Banks et al. 2019), a longitudinal biennial survey representative of individuals aged 50 and over in private households. During the pandemic, ELSA members were invited to participate online or by CATI (Computer-Assisted Telephone Interviewing) to two COVID-19 sub-studies in June/July 2020 and in November/December (75% response rate in both waves, 94% longitudinal response rate). For this study, we restricted our sample to 2,468 grandparents who participated in the second COVID-19 wave (when information on the provision of grandchild care throughout the pandemic was collected); with at least one grandchild aged 15 or younger (as questions on grandparenting were not asked to grandparents with older grandchildren); no missing data on the provision of grandchild care during the pandemic; and with available socio-economic and health information in the most recent pre-pandemic data (wave 9, collected in 2018/19). Further details of the survey’s sampling frame and methodology can be found at www.elsa-project.ac.uk. ELSA was approved by the London Multicentre Research Ethics Committee (MREC/01/2/91), with the COVID-19 sub-studies approved by the UCL REC. Informed consent was obtained from all participants. All data are available through the UK Data Service (SN 8688 and 5050).

### Main measurements of interest

#### Grandparental childcare provision

Information on provision of grandchild care throughout the pandemic was only collected in the second Covid-19 sub-study in November/December 2020. All grandparents with at least one grandchild aged 15 or younger were asked if in February 2020 -before lockdown started-they looked after any of their grandchildren without their parents being present. Those who reported to have looked after grandchildren were then asked whether the amount of care provided to grandchildren changed during the lockdown period (March to June 2020), with options including ‘it increased’, ‘it decreased’, ‘it stayed the same’, or ‘it stopped’. Similar questions were also asked about changes in grandchild care provision during the summer months (June, July, and August 2020), and during the months since schools re-opened (September, October, November/ December 2020) compared to pre-pandemic levels. Based on the possible combinations of answers, we classified respondents into four broad categories distinguishing between (i) those who did not provide grandchild care pre-pandemic; (ii) those who during the first 8 – 9 months of the pandemic stopped completely to look after grandchildren; (iii) those who overall decreased or interrupted their grandchild care; and (iv) those who reported mostly the same or an increase in the amount of time they looked after grandchildren. Full details about all possible patterns of changes and how grandparents were classified can be found in Supplementary Table S1.

#### Mental Health

We considered three outcome measures of mental health assessed at the second COVID-19 sub-study (Nov/Dec 2020): depressive symptoms, quality of life, and life satisfaction. Symptoms of depression were measured by an abbreviated version of the validated Centre for Epidemiologic Studies Depression (CES-D) Scale. The CES-D scale is not a diagnostic instrument for clinical depression but can be used to identify people ‘at risk’ of depression in population-based studies (Radloff 1977). This short version has good internal consistency (Cronbach’s α >0.95) and comparable psychometric properties to the full 20-item CES-D. The scale includes 8 binary (no/yes) questions that enquire about whether respondents experienced any depressive symptoms, such as feeling sad or having restless sleep, in the week prior to the interview. We classified respondents who reported four or more depressive symptoms on the CES-D scale as with elevated depressive symptoms. Furthermore, we considered subjective quality of life (QoL) evaluated using the CASP-12 scale. This is an abbreviated measure of the validated CASP-19 scale which was specifically designed for individuals in later life and used in a wide variety of ageing surveys (Hyde et al. 2003). CASP-12 contains 12 Likert-scaled questions measuring older people’s control and autonomy as well as self-realization through pleasurable activities. The possible range of CASP-12 scores is from 0 to 36, with higher scores indicating greater well-being; CASP-19 is treated as a continuous variable. Finally, we considered life satisfaction as a measure of personal well-being assessed using the Office for National Statistics (ONS) well-being scale (“On a scale of 0 to 10, where 0 is “not at all” and 10 is “very”, how satisfied are you with your life nowadays?”). This allows respondents to integrate and weigh various life domains the way they choose (Pavot and Diener 1993).

#### Covariates

Several potential confounders related to demographic characteristics, socio-economic circumstances, social support, and family characteristics were adjusted for in all multivariate analyses. The majority of these variables are known to be associated with grandparental childcare provision (Di Gessa et al. 2016a; Hank and Buber 2009; Igel and Szyklik 2011) as well as with the main dependent variables under study (Vink et al. 2008; Webb et al. 2011). We controlled for age and age squared to account for non-linear relationships with the outcome variables; sex; and ethnicity (White vs non-White participants due to data constraints in ELSA). To capture respondents’ socio-economic characteristics we controlled for pre-pandemic education, income, wealth, housing tenure, and paid employment (the latter during the pandemic). Educational level was recoded into low (below secondary), middle, and high (university or above) following the International Standard Classification of Education (http://www.uis.unesco.org/). We categorised respondents by quintiles of wealth (total net non-pension non-housing wealth) and accounted for their equivalised total income (from paid work, state benefits, pensions and assets). Housing tenure distinguished outright owners, owners with a mortgage, and non-owners. Paid employment distinguished retired, in paid work and not working from home, in paid work and mostly working from home, furloughed, and other (including homemakers, unemployed, and sick or disabled).

We also accounted for pre-pandemic health and social support during the pandemic. In particular, we controlled for disability (having impairments with basic and instrumental activities of daily living) and clinical vulnerability to COVID-19 (defined irrespective of age as reporting chronic lung disease, asthma, coronary heart disease, Parkinson’s disease, multiple sclerosis, diabetes; weakened immune system as a result of cancer treatment in the previous two years; BMI of 40 or above; or having been advised to shield by their GP/NHS) (Di Gessa and Price 2021a; UK GOV 2020). We further controlled for pre-pandemic measures of mental health (see above for derivation). Moreover, we included three measures of social isolation and social support during the pandemic: household composition, social contacts, and loneliness. For household composition, we distinguished between respondents living alone, with a partner only, with partner and child(ren), with child(ren) but not the partner, and any other arrangements. For social contacts, we categorised respondents as having infrequent contact if they reported real-time contact (by telephone or video calling) with family outside the household and friends less than once a week or never throughout the pandemic (=0 otherwise). Loneliness was assessed using the short version of the revised UCLA loneliness scale with scores of 6 and higher indicating greater loneliness. Furthermore, we controlled for whether respondents had any friends or family members who were hospitalised or died because of Covid-19 as this could be a proxy of heightened perceived fear of Covid-19 which might affect both mental health and the likelihood of wanting to mix with family members and friends outside the household.

Finally, as family structures have been associated with the provision of grandparental childcare (Di Gessa et al. 2016a; Hank and Buber 2009; Igel and Szyklik 2011), we included the following grandchildren’s characteristics: total number of grandchildren; time to travel to their nearest grandchild (living in the same household or less than 15 minutes away; between 15 and 30 minutes away; more than 30 minutes away); and age of the youngest grandchild distinguishing between 0 to 2, 3 to 5, and 6 to 15 years. However, it is worth mentioning that these covariates were obtained from data collected in Wave 9, and that the distance to the closest grandchild and the age of the youngest one do not necessarily refer to the grandchild grandparents were looking after before the pandemic.

### Statistical Analysis

Following descriptive analysis, we investigated the longitudinal associations between patterns of grandparental childcare provision and mental health using logistic or linear models depending on the outcome. For all analyses, we present fully adjusted results accounting for socio-demographic, economic, and health covariates (including pre-pandemic relevant mental health measure) and grandchildren’s characteristics. All analyses were performed using Stata 16. Longitudinal sampling weights were employed to account for different probabilities of being included in the sample and for nonresponse to the survey.

## Results

### Descriptive statistics

Table 1 shows socio-economic, health, and demographic characteristics of grandparents with at least one grandchild aged 15 and younger by whether they were providing grandchild care pre-pandemic. Overall, 52% of grandparents were looking after grandchildren in February 2020. Grandparents who provided any grandchild care pre-pandemic were more likely to be younger, female, and to report better physical health. Looking after grandchildren pre-pandemic is also associated with more advantaged characteristics such as being in paid work, being a homeowner mortgage-free, and being in the highest wealth quintiles. The distribution of the social isolation and support suggests few differences between grandparents who looked after grandchildren and those who did not, with a higher percentage of grandparents who did not provide pre-pandemic care reporting infrequent contacts during the first 8-9 months of the pandemic.

**Table 1.**
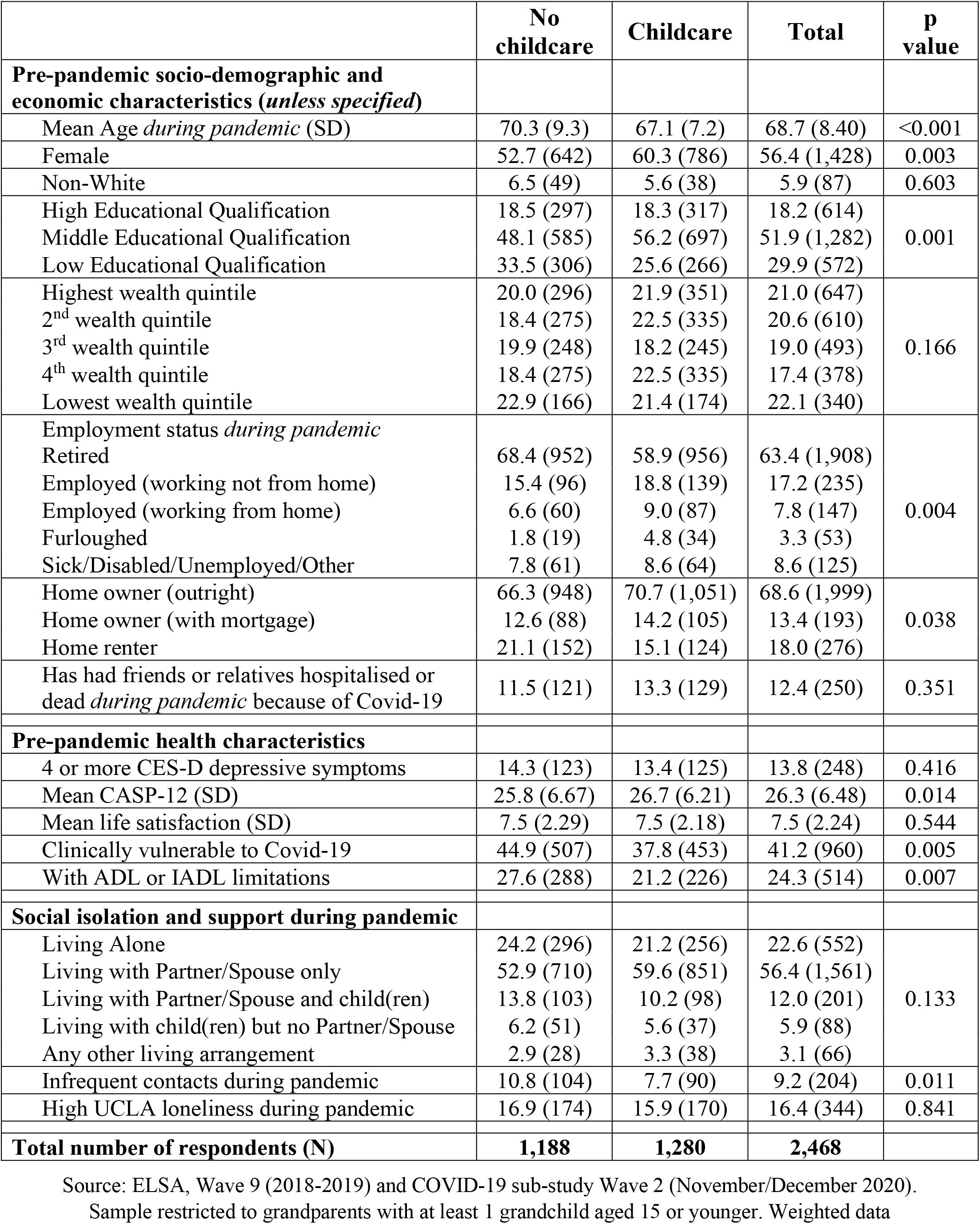
Percent distribution (N) of socio-demographic, economic, health, and social contact characteristics, by pre-pandemic grandchild care provision.

Figure 1 shows that overall, among grandparents who provided grandchild care pre-pandemic, the percentage of ELSA grandparents who completely stopped looking after them was highest during the initial months of the pandemic (53%) when a national lockdown was announced and people were advised to stay at home as much as possible and to minimise contacts outside of their households. However, even during the summer of 2020 when some restrictions were lifted, more than a quarter of grandparents (28%) reported stopping caring for their grandchildren, with a higher percentage in the autumn (38%) when infections rates in England increased again and new restrictions were introduced. Similarly, between 20 and 30% of grandparents reported that the amount of care they provided for their grandchildren decreased compared to pre-pandemic. Grandparents who reported that the amount of grandchild care provided increased during the pandemic were around 6-7% when restrictions were in place (spring and autumn 2020) and about 10% over the summer.

**Figure 1.**
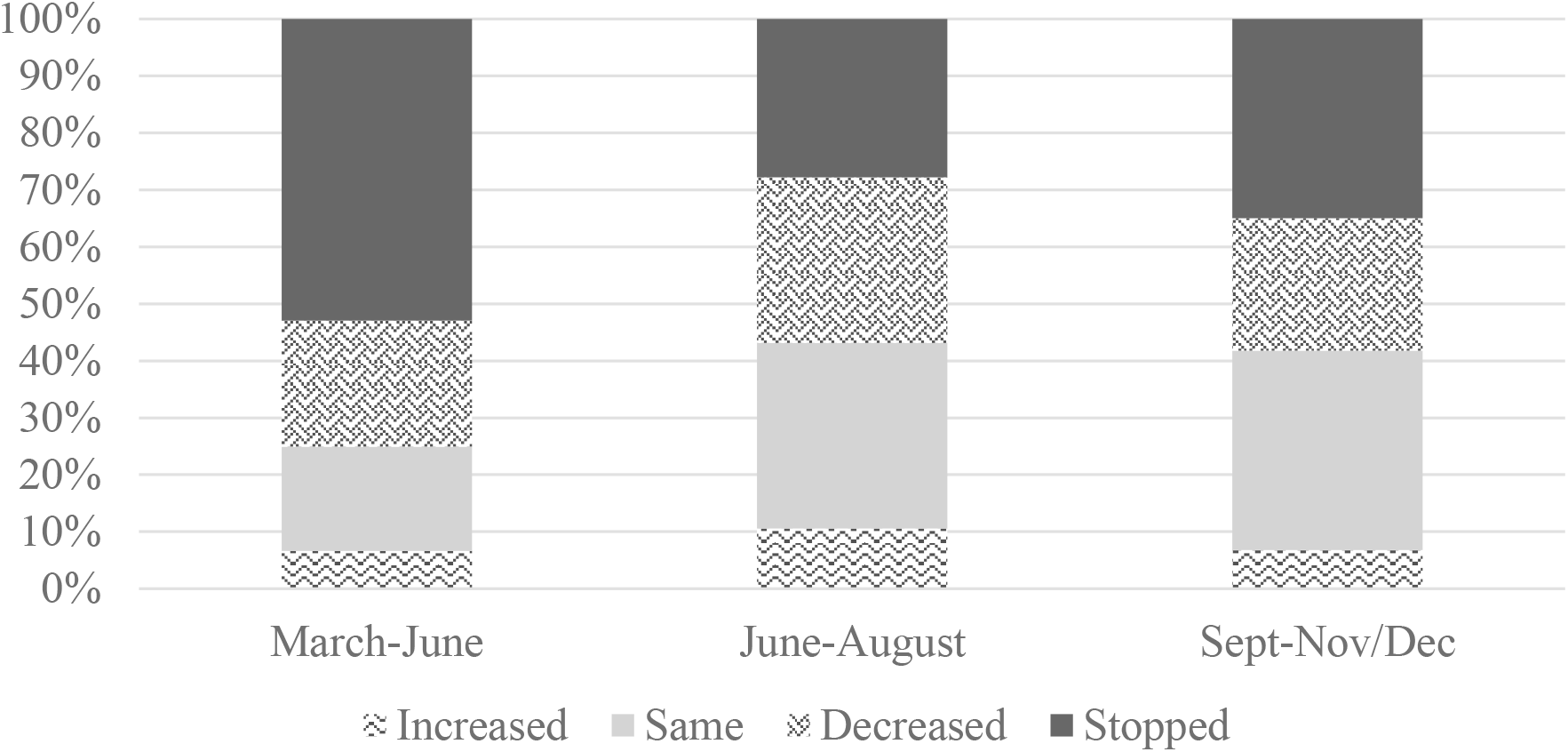
Changes over time in the amount of grandparental childcare provision compared to pre-pandemic levels. Source: ELSA COVID-19 sub-study Wave 2 (November/December 2020). Weighted data. Analyses restricted to grandparents with grandchildren aged 15 or younger who provided care pre-pandemic in February 2020.

Table 2 shows the patterns of grandparental childcare over the first 8-9 months of the pandemic. Just less than half of the grandparents reported that they were not looking after grandchildren in February 2020 (before the pandemic). About one in five grandparents reported that the time spent looking after grandchildren was mostly the same or increased compared to pre-pandemic levels, whereas 22.4% reported that their engagement in grandchild care was mostly reduced or interrupted, and 9.5% completely stopped caring for their grandchildren.

**Table 2.** Distribution of patterns of grandparental childcare provision across three time points (March-June 2020, June-August 2020, and September-November/December 2020) and unadjusted mental health by patterns of grandparental childcare provision.

|  | **N** | **%** | **% Elevated depressive symptoms (CES-D)** | **Mean quality of life (CASP-12)** | **Mean life satisfaction** |
| --- | --- | --- | --- | --- | --- |
| No grandchild care pre-pandemic | 1,188 | 47.8 | 29.7 | 24.58 | 6.85 |
| Mostly same or increased | 456 | 20.3 | 25.9 | 25.82 | 7.05 |
| Mostly decreased or interrupted | 568 | 22.4 | 27.2 | 25.37 | 6.91 |
| Completely stopped | 256 | 9.5 | 34.3 | 24.58 | 6.65 |
| **N respondents** | **2468** | **100** | **28.9** | **25.02** | **6.88** |

Source: ELSA COVID-19 sub-study Wave 2 (November/December 2020). Weighted data. A more comprehensive table with detailed patterns of changes in grandchild care provision can be found in Supplementary Table S1.

Table 2 also shows that mental health measured in the second wave of the COVID-19 ELSA sub-study showed substantial variation by patterns of grandparental childcare. Respondents who stopped completely looking after grandchildren reported the highest percentages of elevated depressive symptoms (34.3%), the lowest mean life satisfaction (6.65), and the lowest quality of life among grandparents who looked after grandchildren pre-pandemic (Mean CASP-12=24.58). Grandparents who overall had the same or increased levels of engagement in childcare, however, reported the best mental health (with 25.9% reporting elevated depressive symptoms; mean CASP-12 quality of life 25.82; and mean life satisfaction 7.05).

### Multivariable analyses

To investigate how patterns of grandchild care provision during the pandemic were associated with mental health we used logistic and linear regressions, depending on the outcome variable. We present (Table 3) results for the main variable of interest from fully adjusted models (Full results of the fully adjusted models in Supplementary Table S2). Accounting for socio-demographic and economic characteristics as well as for the health profile of the grandparents and their social support during the pandemic, we found significant differences in mental health by patterns of changes in the provision of grandparental childcare. In particular, compared to grandparents who maintained unchanged or increased grandchild care provision during the pandemic, grandparents who stopped completely looking after grandchildren were more likely to report high depressive symptoms (OR=1.91; 95% CI = 1.17 to 3.11), lower levels of quality of life (B=-1.17, 95% CI= -2.02 to -0.32), and a marginal statistical significant (p=0.066) lower life satisfaction (B=-0.36; 95% CI = -0.74 to 0.02). Analyses also suggest that compared to those who maintained grandchild care during the pandemic, also those who mostly reduced or interrupted their provision of childcare had lower levels of quality of life (B=-0.76; 95% CI = -1.39 to -0.13) and lower life satisfaction (B=-0.28; 95% CI= - 0.56 to -0.01). The direction of the association is similar also for decrease in grandchild care and depression, although this was not statistically significant.

**Table 3.** Associations between grandparental childcare patterns and mental health. Results from fully-adjusted logistic and linear regression models.

|  | **Elevated depressive symptoms (CES-D)** | **Quality of Life (CASP-12)** | **Life Satisfaction** |
| --- | --- | --- | --- |
| No grandchild care pre-pandemic | 1.37<br>[0.93,2.01] | -0.80**<br>[-1.37,-0.23] | -0.21<br>[-0.48,0.06] |
| Mostly same or increased | *Ref* | *Ref* | *Ref* |
| Mostly decreased or interrupted | 1.25<br>[0.81,1.92] | -0.76*<br>[-1.39,-0.13] | -0.28*<br>[-0.56,-0.01] |
| Completely stopped | 1.91**<br>[1.17,3.11] | -1.17**<br>[-2.02,-0.32] | -0.36+<br>[-0.74,0.02] |
| *N respondents* | 2,427 | 2,297 | 2,265 |

Sources: ELSA, COVID-19 sub-study Wave 2 (November/December 2020) and Wave 9 (2018/19). Notes: Fully-adjusted models adjusted for age, age squared, sex, ethnicity, education, income, wealth, home tenure, employment, pre-pandemic disability, clinical vulnerability to COVID-19, relevant pre-pandemic mental health variable, household composition, social contacts, loneliness, number of grandchildren, distance to the closest grandchild, age of the youngest grandchild, as well as an indicator of whether friends and family members got hospitalised or died of Covid-19. Odds Ratios [and 95% confidence intervals (CIs)] reported for elevated depressive symptoms, and Beta coefficients [and 95% CIs] for the continuous outcome variables ‘Quality of life’ and ‘Life Satisfaction’. For both continuous outcomes, the relevant health questions in Wave 9 were asked in the self-completion questionnaire. Analyses are restricted to grandparents who reported grandparental childcare pre-pandemic. + $p < 0.10$ , \* $p < 0.05$ , \*\* $p < 0.01$ , \*\*\* $p < 0.001$ . Weighted data. Detailed models can be found in the supplementary Tables S2.

### Robustness Checks

Given that the category of grandparents who mostly decreased or interrupted their childcare provision is heterogeneous as it also includes some grandparents who, for one of the three time points considered, reported the same or increased levels of childcare provision, we conducted some checks to test the robustness of our results. In particular, we used a stricter definition of ‘decrease/interruption’ of grandchild care provision, and further distinguished grandparents who reported that their amount of care ‘decreased’ or ‘stopped’ *at all three periods considered*. The results obtained with this classification of changes in grandparental childcare were broadly similar to the ones reported above (as shown in Supplementary Table S3), with almost identical coefficients and similar strength of associations for both those who ‘mostly’ and those who ‘only’ decreased or interrupted childcare. However, the strength of these associations is somehow weaker, most likely because of the smaller sample size in each of these two categories.

## Discussion

Providing grandparental childcare was quite common pre-pandemic. However, during the pandemic, grandparents were strongly advised to stop looking after grandchildren and more generally to limit their physical interactions with younger people, despite the positive impact that intergenerational relationships have on mental health. In this paper, therefore, we investigate the impact of changes in grandparental provision during the pandemic on three measures of mental health, namely depression, quality of life, and life satisfaction among grandparents in England.

Our analyses showed that overall, compared to grandparents who maintained unchanged or increased grandchild care provision during the pandemic, those who stopped looking after grandchildren altogether during the first 8/9 months of the pandemic as well as those who mostly reduced their involvement in grandchild care reported generally poorer mental health. Therefore, this study supports the idea that abrupt interruptions of older people’s role in society and family (in this study, of grandparents’ roles as care providers) have negative consequences for their mental health, in line with the ‘ambiguous loss’ argument and studies that found that grandparents unable to see and spend time with their grandchildren for reasons beyond their control might get frustrated and distressed about it, with negative consequences for their mental health (Drew and Smith 1999; Drew and Silverstein 2007). The COVID-19 pandemic might have acted in a very similar way, given that for many grandparents the pandemic, over which they could not exert any control, meant stopping or disrupting the amount of time spent looking after grandchildren (because of the guidelines or personal/family fears to get together) without knowing when they could be able to spend time with their grandchildren the same way they used to pre-pandemic.

### Strengths and limitations

We investigated associations between changes in grandparental childcare provision and grandparents’ mental health during the pandemic. To our knowledge, this was the first study to investigate this issue drawing strength from using longitudinal data from the nationally-representative English Longitudinal Study of Ageing. Our analysis supports the idea that both stopping altogether and disruptions to grandchild care provision have had an additional detrimental effect on grandparent’s mental health, perhaps because of the lack of control over this interruption of care (dictated by the pandemic itself and government responses that are largely beyond people’s control) combined with the uncertainty of not knowing how long until they could spend time with them again.

Our contribution, however, should be considered in light of some limitations. First, as mentioned above, ELSA does not collect detailed information about the childcare provided to each grandchild, but rather asks a more generic question related to all grandchildren and changes in the amount of time spent looking after them. Moreover, although in our analyses we considered several grandchildren’s characteristics (such as the age of the youngest grandchild and where the nearest grandchild lives), this information was collected in the pre-pandemic Wave 9 and might not relate to the grandchild to which grandparents provide care. Second, although the intergenerational decision-making process is generally related to the opportunities and resources of all three generations (Price et al. 2018), and particularly during the pandemic might have reflected both parents’ employment status as well as school closures and (lack of) availability of formal childcare (Bordone et al. 2016; Di Gessa et al. 2016a; Igel and Szyklik 2011), ELSA does not collect any information on parents and on any childcare arrangements. Third, although in our analyses we controlled for whether grandparents had relatives or friends severely affected by COVID-19, ELSA did not collect information about respondents’ perception on individuals’ ability to tolerate and cope with the uncertainty due to COVID-19, or personality characteristics such as degree of risk tolerance or harm avoidance. These factors might help further understand both different behaviours and choices around levels of and changes in grandparental childcare and their subsequent effect on mental health. Fourth, although we controlled for social interactions during the pandemic, we did not have any specific measure on non-physical contacts specifically with grandchildren. Fifth, we only had information about the provision of grandchild care collected in one Wave, referring to three broad points in time, and only referring to changes without detailing the nuanced frequency of childcare provision pre and during the pandemic. While we cannot construct more nuanced measures of grandchild care provision, this is likely to be the best data obtainable at scale for this activity among grandparents during the pandemic. Sixth, ELSA suffers from non-random cumulative attrition, an unavoidable problem in longitudinal studies which can only partially be corrected by using weights in the analysis. Finally, although ELSA is limited to the population of England, we believe that we can draw inferences from this study also for (at least) other European countries where policy recommendations and media advice were, in the early phases of the pandemic, quite similar and all pointed towards limiting as much as possible interactions between grandparents and grandchildren. If anything, we can expect that the negative associations observed between interruptions/disruptions of grandchild care and grandparents’ mental health would be larger in those countries (e.g., Mediterranean countries) where grandparental childcare is more normative, formal childcare services are limited, and grandparents tend to provide more regular grandchild care (Bordone et al. 2016; Di Gessa et al. 2016a; Igel and Szyklik 2011).

In summary, our study provides a picture of the broader consequences of the pandemic among a considerable proportion of older people, i.e. grandparents who changed the amount of time they looked after grandchildren. Although recommendations to stop caring for grandchildren had the clear aim to save grandparents’ lives, it should be acknowledged that such disruption had adverse consequences for the mental health of both grandparents who stopped altogether looking after grandchildren and those who mostly reduced the amount of grandchild care provided. In emerging from the current pandemic and if physical distancing policies remain a core strategy to protect individuals at higher risk from COVID-19 variants or indeed in a future pandemic, attention should be paid to addressing the mental health and wider needs of older people who may suffer from the loss of their roles.

## Data Availability

All data produced are available online from the UK Data Service

https://beta.ukdataservice.ac.uk/datacatalogue/studies/study?id=8688

https://beta.ukdataservice.ac.uk/datacatalogue/studies/study?id=5050

## Declarations

### Availability of data and material

Researchers can download ELSA data from all waves, from the UK Data Service. For more information, please visit https://www.elsa-project.ac.uk/accessing-elsa-data

### Authors’ contribution

G Di Gessa conducted analyses, drafted the paper, and co-led the interpretation of data with B Arpino and V Bordone. G Di Gessa and V Bordone led the conception and design of the study. B Arpino contributed to the study design. All authors contributed to the interpretation of data and manuscript revisions. All authors have read and approved the final version.

## Acknowledgements

The data were made available through the UK Data Archive. The English Longitudinal Study of Ageing was developed by a team of researchers based at University College London, NatCen Social Research, the Institute for Fiscal Studies, the University of Manchester and the University of East Anglia. The data were collected by NatCen Social Research. The funding is currently provided by the National Institute on Aging in the US, and a consortium of UK government departments coordinated by the National Institute for Health Research. Funding has also been received by the Economic and Social Research Council.

