## Supplementary Table for "Grandparental Childcare During the Pandemic and Effects on Mental Health: Evidence From England"

*Supplementary Table S1. Detailed patterns of changes in provision of grandchild care during the pandemic*

| Classification of changes in amount of childcare | Lockdown (March-June) | Summer (June-August) | Autumn (Sept-Nov/Dec) | N | Unweighted percentage |
|---|---|---|---|---|---|
| **Mostly same or increased** | Same/Increased | Same/Increased | Same/Increased | 189 | 7.66 |
| | Same/Increased | Same/Increased | Decreased | 34 | 1.38 |
| | Same/Increased | Same/Increased | Stopped | 12 | 0.49 |
| | Same/Increased | Decreased | Same/Increased | 14 | 0.57 |
| | Decreased | Same/Increased | Same/Increased | 56 | 2.27 |
| | Stopped | Same/Increased | Same/Increased | 136 | 5.51 |
| | Same/Increased | Stopped | Same/Increased | 15 | 0.61 |
| **Mostly decreased or interrupted** | Same/Increased | Decreased | Decreased | 14 | 0.57 |
| | Same/Increased | Decreased | Stopped | 2 | 0.08 |
| | Same/Increased | Stopped | Decreased | 2 | 0.08 |
| | Same/Increased | Stopped | Stopped | 9 | 0.36 |
| | Decreased | Same/Increased | Decreased | 31 | 1.26 |
| | Decreased | Same/Increased | Stopped | 4 | 0.16 |
| | Decreased | Decreased | Same/Increased | 30 | 1.22 |
| | Decreased | Decreased | Decreased | 105 | 4.25 |
| | Decreased | Decreased | Stopped | 18 | 0.73 |
| | Decreased | Stopped | Same/Increased | 4 | 0.16 |
| | Decreased | Stopped | Decreased | 6 | 0.24 |
| | Decreased | Stopped | Stopped | 11 | 0.45 |
| | Stopped | Same/Increased | Decreased | 32 | 1.30 |
| | Stopped | Same/Increased | Stopped | 24 | 0.97 |
| | Stopped | Decreased | Same/Increased | 54 | 2.19 |
| | Stopped | Decreased | Decreased | 74 | 3.00 |
| | Stopped | Decreased | Stopped | 66 | 2.67 |
| | Stopped | Stopped | Same/Increased | 53 | 2.15 |
| | Stopped | Stopped | Decreased | 29 | 1.18 |
| **Completely stopped** | Stopped | Stopped | Stopped | 256 | 10.37 |
| **No grandchild care pre-pandemic in Feb 2020** | -- | -- | -- | 1,188 | 48.14 |

Source: ELSA, COVID-19 sub-study Wave 2 (November/December 2020). Columns 2-4 specify whether the respondents reported increases, decreases, no changes, or interruption of grandchild care provision at each time point. Unweighted data

***Supplementary Table S2. Associations between grandparental childcare patterns and mental health and well-being. Fully-adjusted regression models.***

| | Elevated depressive symptoms (CES-D) | Quality of Life (CASP-12) | Life Satisfaction |
|---|---|---|---|
| No grandchild care pre-pandemic | 1.37 [0.93,2.01] | -0.800** [-1.37,-0.23] | -0.209 [-0.48,0.06] |
| Mostly same or increased | *Ref* | *Ref* | *Ref* |
| Mostly decreased or interrupted | 1.25 [0.81,1.92] | -0.761* [-1.39,-0.13] | -0.282* [-0.56,-0.01] |
| Completely stopped | 1.91** [1.17,3.11] | -1.173** [-2.02,-0.32] | -0.359+ [-0.74,0.02] |
| Female (Ref: Male) | 1.50** [1.12,2.00] | -0.613** [-1.04,-0.19] | -0.275** [-0.46,-0.09] |
| Age | 0.98 [0.96,1.01] | 0.013 [-0.03,0.05] | 0.019* [0.00,0.04] |
| Age squared | 1.00 [1.00,1.00] | -0.004** [-0.01,-0.00] | -0.002 [-0.00,0.00] |
| Non-White (Ref: White) | 0.84 [0.42,1.67] | 1.402 [-0.22,3.02] | 0.344 [-0.21,0.90] |
| Education (Ref: Low) | | | |
| Medium education | 1.00 [0.70,1.43] | 0.240 [-0.25,0.73] | 0.260* [0.05,0.47] |
| Low education | 1.13 [0.74,1.72] | 0.436 [-0.23,1.11] | 0.350* [0.08,0.62] |
| Wealth (Ref: Lowest quintile) | | | |
| 2nd lowest quintile | 0.89 [0.56,1.42] | 0.472 [-0.37,1.32] | -0.207 [-0.61,0.19] |
| 3rd wealth quintile | 0.86 [0.53,1.40] | 0.387 [-0.49,1.26] | -0.0805 [-0.46,0.30] |
| 4th wealth quintile | 0.69 [0.42,1.12] | 0.486 [-0.36,1.33] | -0.103 [-0.48,0.28] |
| Highest wealth quintile | 0.60 [0.35,1.02] | 0.486 [-0.40,1.37] | -0.204 [-0.60,0.19] |
| Income | 1.00 [0.91,1.09] | 0.0141 [-0.07,0.10] | -0.00284 [-0.04,0.03] |
| Employment status (Ref: Retired) | | | |
| Employed not Working From Home (WFH) | 0.45** [0.25,0.82] | 1.065** [0.36,1.77] | 0.439** [0.12,0.76] |
| Employed mostly WFH | 1.52 [0.89,2.60] | 0.829 [-0.02,1.68] | 0.260 [-0.10,0.63] |
| Furloughed | 1.10 [0.51,2.39] | 0.534 [-1.13,2.20] | 0.519 [-0.24,1.28] |
| Other employment | 1.42 [0.79,2.53] | 0.430 [-0.70,1.56] | 0.286 [-0.23,0.81] |
| Housing (Ref: mortgage-free owner) | | | |
| Homeowner with mortgage | 1.25 [0.74,2.11] | -0.0896 [-0.89,0.71] | -0.123 [-0.49,0.24] |
| Rented accommodation | 0.78 [0.48,1.26] | 0.206 [-0.65,1.06] | 0.0643 [-0.29,0.42] |
| Clinically vulnerable to COVID-19 | 1.09 [0.82,1.46] | -0.286 [-0.71,0.14] | 0.0417 [-0.16,0.24] |
| Disability | 2.14*** [1.55,2.95] | -1.450*** [-2.10,-0.80] | -0.576*** [-0.85,-0.30] |
| Distance (Ref: ≤15m or cohabiting) | | | |
| Between 15 and 30m | 1.30 [0.92,1.83] | -0.105 [-0.62,0.41] | -0.296* [-0.53,-0.07] |
| More than 30m | 0.99 [0.70,1.40] | 0.279 [-0.21,0.77] | -0.290* [-0.52,-0.06] |
| Number of grandchildren | 1.01 [0.98,1.04] | 0.0555 [-0.01,0.12] | 0.0219 [-0.00,0.05] |
| Age youngest grandchild (Ref: 0-2) | | | |
| 3-5 | 0.89 [0.61,1.29] | 0.669* [0.08,1.26] | 0.0769 [-0.17,0.33] |
| 6-15 | 0.81 [0.58,1.12] | 0.110 [-0.41,0.63] | 0.00592 [-0.22,0.24] |
| Living arrangements (Ref: partnered) | | | |
| Living Alone | 1.17 | 0.497 | 0.106 |

| | | | |
|---|---|---|---|
| | [0.83,1.63] | [-0.05,1.05] | [-0.14,0.35] |
| Living w\ partner & children | 1.00<br>[0.62,1.60] | -0.701<br>[-1.45,0.05] | -0.168<br>[-0.47,0.14] |
| Single parent | 1.74<br>[0.88,3.45] | -0.185<br>[-1.53,1.16] | 0.0301<br>[-0.53,0.59] |
| Other living arrangements | 1.55<br>[0.63,3.83] | -1.053*<br>[-2.07,-0.04] | 0.0346<br>[-0.48,0.55] |
| Infrequent contacts with friends and family throughout pandemic | 1.00<br>[0.61,1.64] | -0.699<br>[-1.44,0.04] | -0.0407<br>[-0.45,0.37] |
| High loneliness at one Covid Wave | 4.73***<br>[3.41,6.56] | -3.136***<br>[-3.77,-2.50] | -1.137***<br>[-1.41,-0.86] |
| High loneliness throughout pandemic | 12.34***<br>[8.71,17.48] | -4.758***<br>[-5.52,-3.99] | -1.592***<br>[-1.93,-1.25] |
| Has had friends/relatives hospitalised or dead because of Covid-19 | 1.50*<br>[1.02,2.20] | -0.172<br>[-0.96,0.61] | -0.242<br>[-0.56,0.08] |
| Pre-pandemic measure of relevant mental health/ well-being | 3.69***<br>[2.39,5.71] | 12.04***<br>[10.22,13.87] | 5.209***<br>[4.51,5.91] |
| Observations | 2427 | 2297 | 2265 |

Sources: ELSA, COVID-19 sub-study Wave 2 (November/December 2020), COVID-19 sub-study Wave 1 (June/July 2020) and Wave 9 (2018/19). Notes: Odds Ratios [and 95% CIs] reported for elevated depressive symptoms, and Beta coefficients [and 95% CIs] for the continuous outcome variables ‘Quality of life’ and ‘Life Satisfaction’. For both continous outcomes, the relevant health questions in Wave 9 were asked in the self-completion questionnaire. Analyses are restricted to grandparents who reported grandparental childcare pre-pandemic.* $p < 0.05$, ** $p < 0.01$, *** $p < 0.001$. Weighted data.

***Supplementary Table S3. Robustness checks of the associations between grandparental childcare patterns and mental health and well-being using a stricter classification of 'decrease/interruption'. Fully-adjusted regression models.***

| | **% (N)** | **Elevated depressive symptoms (CES-D)** | **Quality of Life (CASP-12)** | **Life Satisfaction** |
|---|---|---|---|---|
| No grandchild care pre-pandemic | 47.8 (1188) | 1.37 [0.93,2.01] | -0.800** [-1.37,-0.23] | -0.209 [-0.48,0.06] |
| Mostly same or increased | *20.3 (456)* | *Ref* | *Ref* | *Ref* |
| Only decreased or interrupted | 12.4 (309) | 1.36 [0.85,2.16] | -0.774* [-1.54,-0.01] | -0.296+ [-0.63,0.04] |
| Mostly decreased or interrupted | 10.0 (259) | 1.12 [0.63,2.01] | -0.745* [-1.50;-0.00] | -0.265+ [-0.57,0.04] |
| Completely stopped | 9.5 (256) | 1.91** [1.17,3.11] | -1.173** [-2.02,-0.33] | -0.359+ [-0.74,0.02] |
| *N respondents* | *2,468* | *2,427* | *2,297* | *2,265* |

Sources: ELSA, COVID-19 sub-study Wave 2 (November/December 2020) and Wave 9 (2018/19). Notes: Fully-adjusted models adjusted for age, age squared, sex, ethnicity, education, income, wealth, home tenure, employment, pre-pandemic disability, clinical vulnerability to COVID-19, relevant pre-pandemic mental health variable, household composition, social contacts, loneliness, number of grandchildren, distance to the closest grandchild, age of the youngest grandchild, as well as an indicator of whether friends and family members got hospidalised or died of Covid-19. Odds Ratios [and 95% confidence intervals (CIs)] reported for elevated depressive symptoms, and Beta coefficients [and 95% CIs] for the continuous outcome variables 'Quality of life' and 'Life Satisfaction'. For both continous outcomes, the relevant health questions in Wave 9 were asked in the self-completion questionnaire. Analyses are restricted to grandparents who reported grandparental childcare pre-pandemic.+ p0.10, * p < 0.05, ** p < 0.01, *** p < 0.001.
